# Colon-delivered vitamin B2 as a functional modulator of the human gut microbiome

**DOI:** 10.64898/2026.04.01.26349391

**Authors:** Robert E. Steinert, Wilbert Sybesma, Ali May, Chengyao Peng, Thomas Abeel, Pernille Neve Myers, Liang Wu, Floris Klein Obbink, Emiel Ver Loren van Themaat, Jonas Wittwer Schegg, Jérôme Wojcik, Ateequr Rehman

## Abstract

Vitamin B2 (riboflavin) is a key redox cofactor that may modulate gut microbial ecology, yet conventional supplements are absorbed proximally and have limited colonic exposure. We evaluated whether colon-targeted riboflavin alters microbiome composition, function and network structure as well as host biomarkers in healthy older adults. In a randomized, double-blind, placebo-controlled, parallel-group clinical trial (N=348; 50–70y), participants received colon-targeted riboflavin (1.4, 10, or 75 mg/day) or placebo for 12 weeks. The study’s primary endpoint was the change in fecal microbial composition while secondary endpoints encompassed microbiome function, host health biomarkers, and clinical outcomes. Shotgun metagenomics and fecal/blood biomarkers were assessed at baseline, week 4, and week 12. Although no significant changes were observed between groups in overall community-wide diversity metrics (alpha and beta diversity), we found that colon-delivered riboflavin significantly altered the relative abundance of several microbial taxa when compared with placebo. The most pronounced effects on microbiome composition, function, and network structure were observed with the 10 mg dose at week 12, reflected by within group increases in alpha diversity, the largest rise in total species counts, higher HACK index values indicating greater community resilience, and distinct shifts in KEGG module abundance, including enhanced potential for riboflavin biosynthesis. Supplementation with 75 mg riboflavin led to higher fecal butyrate concentrations at week 4 versus placebo, while the lowest dose (1.4 mg) significantly reduced the dysbiosis index within groups and modestly improved network structure across groups. All three doses (1.4, 10, and 75 mg) influenced keystone species abundance. No between-group differences were observed for gastrointestinal symptoms, quality-of-life measures, fecal pH, high-sensitivity C-reactive protein (hs-CRP), calprotectin or the soluble cluster of differentiation 14 (sCD14), a marker of gut barrier integrity, except an increase in plasma riboflavin concentrations at 75 mg after 12 weeks, indicating colonic absorption. The product was safe and well-tolerated across all doses. Our results indicate that colon-targeted riboflavin can act as a functional modulator of the human gut microbiome with the most consistent effects observed at 10 mg and additional dose-specific effects at 1.4 mg and 75 mg. These findings provide novel evidence that microbiome-directed micronutrients can enhance microbial metabolic resilience and cross-feeding capacity. Future studies are warranted to establish related health benefits, either as a standalone intervention or in combination with classical pre-, pro-, or postbiotics, particularly in target populations such as individuals with IBS, stress, mild cognitive decline, or early metabolic and inflammatory alterations.

## Introduction

The human gut microbiota has emerged as a pivotal factor for human health, with mounting evidence linking dysbiosis (defined as an imbalance in microbial composition) to numerous conditions such as obesity, type 2 diabetes, inflammatory bowel diseases (IBD) or neurological disorders (1–4). While conventional approaches such as pre- and probiotics have been widely studied for their microbiome-modulating properties, the role of vitamins remains less explored despite growing evidence of their influence on microbial ecology. In fact, recent research highlights B-vitamins to serve as key metabolic cofactors that are commonly exchanged between microbes for energy production, detoxification, and protection against oxidative stress (5–8). The so called “pantryome model” emphasizes the networked nature of microbial communities, where interactions and shared micronutrients support ecosystem function by sustaining microbial stability through common metabolic resources (5,9–11). This mechanism expands upon the classical model of cross-feeding, which primarily considers microbial interactions as sequential metabolic exchanges involving compounds like lactate, acetate, or other macromolecule degradation products. Beyond carbon and nitrogen-based interactions, it is increasingly evident that micronutrient cross-feeding, including the sharing of B-vitamins, may also play a significant role in shaping microbial community dynamics.

Vitamin B2 (riboflavin) is an essential water-soluble vitamin that serves as a biochemical precursor for the enzymatic cofactors flavin mononucleotide (FMN) and flavin adenine dinucleotide (FAD). Flavin cofactors are indispensable for numerous redox reactions that drive cellular metabolism, supporting both growth and differentiation in all forms of life, including bacteria, archaea and eukarya (8,12–14). In addition, riboflavin-dependent enzymes play key roles in antioxidant defense, given FMN and FAD are co-enzymes required by glutathione reductase and peroxidase, superoxide dismutase and catalase, which all protect cells from the harmful effect of reactive oxygen species (ROS) (15,16). Finally, riboflavin contributes to flavin-based extracellular electron transfer (FLEET), a process in which certain anaerobic gut bacteria (e.g., lactic acid bacteria and *F. prausnitzii*) export electrons via flavins to external acceptors, thereby enhancing energy conservation (respiration without oxygen as an alternative to fermentation) and mitigating oxidative stress under microaerophilic conditions (17–19). Collectively, it is thought that riboflavin shapes not only host physiology but also contributes to gut microbial ecology, favoring microorganisms that can efficiently produce energy and preserve redox balance, thereby enhancing their growth, persistence, and ecological fitness.

In 2015, Thiele and colleagues (20) analyzed the genomes of 256 common human gut bacteria and found that only approximately 12.5% possessed the complete biosynthetic pathways for all eight B vitamins. Interestingly, their findings further revealed an inverse distribution pattern of B-vitamin synthesis, suggesting metabolic interdependence and symbiotic relationships among gut microbes. Regarding riboflavin, they found that its biosynthesis pathway was largely conserved but absent in most *Actinomycetota* genomes and in approximately half of the butyrate-producing *Bacillota* genomes. Notably, all non-riboflavin-producing members of these phyla carried the *RibU* transporter gene, indicating a dependence on environmental or cross-fed riboflavin. This aligns with the long-standing view that B-vitamin-auxotrophic microbes fulfill their requirements by competing for host-derived dietary vitamins—potentially enhanced by fiber-rich, plant-based foods that modulate vitamin bioavailability and promote their natural delivery to the colon—or by scavenging riboflavin from host tissue leakage. Moreover, it underscores the emerging concept that vitamins serve as key mediators of microbial interactions and signaling within the gut ecosystem.

Importantly, recent findings suggest a decrease in microbial vitamin production in oxidative stress-related conditions including IBD, malnutrition, type 2 diabetes mellitus, obesity and low fiber diets highlighting the possible requirement for enhanced external riboflavin supply (13,21–25).

Preliminary *in-vitro* and clinical studies suggest that riboflavin supplementation beneficially modulates gut microbial activity without substantially altering microbial composition. For example, the RIBOGUT randomized trial demonstrated that high-dose oral riboflavin supplementation of 50 mg/day and 100 mg/day in healthy people for two weeks enhanced fecal butyrate production despite no distinct shifts in the relative abundance of known butyrate-producing genera such as *Faecalibacterium prausnitzii*. Moreover, riboflavin supplementation was associated with increased microbial network complexity and metabolic cross-feeding interactions, indicating enhanced microbial resilience and functionality (26). In the RISE-UP study, three weeks of high dose of oral riboflavin (100 mg/d) in patients with Crohn’s disease (CD) further improved systemic oxidative stress and inflammatory markers including interleukin-2 and C-reactive protein indicating an anti-inflammatory effect (27). These findings align with evidence that riboflavin pathway metabolites can activate mucosal-associated invariant T (MAIT) cells, key players in mucosal immunity contributing to both anti-inflammatory and immunomodulatory responses (28).

Conventional riboflavin supplements are largely absorbed in the proximal small intestine, resulting in limited interaction with the dense microbial communities of the colon. In contrast, colon-targeted delivery systems circumvent upper gastrointestinal absorption and release bioactives directly into the colon, providing a promising strategy to locally modulate the gut microbiota (11,29–31). Consistent with this, recent clinical pilot studies using colon-targeted formulations of vitamins, including riboflavin, have reported increased microbial diversity, compositional shifts in bacterial populations, and elevated short-chain fatty acid (SCFA) production, highlighting the therapeutic potential of this approach (32).

Despite these first promising observations, systematic evaluations of colon-targeted riboflavin supplementation across different dosages remain limited. The present clinical trial aims to address this research gap by assessing the impact of varying doses of colon-delivered riboflavin on gut microbial composition, metabolic activity and network structure and function. By elucidating these dose-dependent interactions, this study seeks to clarify the utility of riboflavin as a targeted microbiome intervention and contribute to a deeper understanding of micronutrient-driven modulation of gut ecology and systemic health.

## Methods

### Study population

The study enrolled 348 healthy men and women aged 50 to 70 years, with a body mass index (BMI) between 18.5 and 30□kg/m². Key exclusion criteria included the presence of significant acute or chronic illness, smoking, history of drug or alcohol abuse (defined as more than two servings per day), pregnancy, antibiotic use within the past three months, major dietary changes during the same period, eating disorders, adherence to vegetarian or vegan diets, use of enemas or dietary supplements such as prebiotics, probiotics, or fiber exceeding 30□g/day prior to baseline and throughout the intervention, and chronic medication use for active gastrointestinal conditions. Baseline demographics in the Intention To Treat (ITT) population are shown in Supplementary Table S1.

### Study design

This trial was designed as a randomized, double-blind, placebo-controlled, parallel-group clinical study evaluating the effects of 12-week colon-targeted riboflavin supplementation at 1.4 mg, 10 mg, and 75 mg daily doses versus placebo. The primary endpoint for efficacy analysis was the change in fecal microbial composition. Secondary endpoints comprised changes in fecal microbial diversity (alpha and beta diversity) and fecal short-chain fatty acid concentrations (acetate, propionate, butyrate) as well as markers of gut barrier integrity (plasma sCD14), inflammation (fecal calprotectin, plasma hs-CRP), and oxidative stress (plasma free thiols). In addition, changes in gastrointestinal symptoms, stool patterns, quality of life, plasma riboflavin, and fecal pH were assessed. Exploratory investigations included gut microbiome health, functional capacity and organization of the gut microbiome network structure (HACK and Dysbiosis index, riboflavin prototrophic fraction, modular microbiome network structure and performance and functional KEGG modules). The full protocol can be accessed at clinicaltrials.gov (ID:NCT05803811, registration date 2023-02-16).

The trial was coordinated and conducted by Atlantia Clinical Trials Ltd. (Cork, Ireland) with participants attending three on-site visits: Visit 1 (screening) at approximately 2 weeks before the start of the study, visit 2 (baseline) at week 0, and visit 3 (end-of-study) at week 12. At each visit, standard physical exams were conducted, including weight, BMI, and vital signs. At visit 1 (screening), participants also gave written consent, underwent eligibility screening, completed the Block Fiber Screener, <30 g/day cutoff, (NutritionQuest, Berkeley, CA, USA) and received a stool collection kit with instructions for collecting and storing a stool sample. In addition, a blood sample was taken for safety assessment. At visit 2 (baseline), when eligibility was reconfirmed, participants returned the stool sample (which was collected and stored at -20°C in home freezers within 24 hours before the visit) and a blood sample was taken for safety profiling and efficacy analysis. Furthermore, participants completed three questionnaires: a 36-item Short Form Health survey questionnaire (SF-36), the Gastrointestinal Symptom Rating Scale (GSRS) and a Food Frequency Questionnaire (FFQ). The randomized participants subsequently received a 12-week investigational product (IP) study supply and were instructed to take one capsule daily at breakfast and maintain a stable diet. At week 4, no visit took place, but only a second fecal sample was collected to additionally assess SCFA gut microbiome composition and metabolic activity. At visit 3 (end-of-study), 12 weeks after the start of the study, participants returned another fecal sample and another blood sample was collected. Participants also completed the FFQ, SF-36 and GSRS. Compliance was evaluated via capsule returns.

### Study product

The investigational products were riboflavin (riboflavin table grade, DSM-Firmenich, Kaiseraugst Switzerland) tested at three different doses (1.4 mg, 10 mg, and 75 mg), or placebo (microcrystalline cellulose, VIVAPUR 102 JRS Pharma GmbH & Co KG, Rosenberg Germany), each administered once daily in the form of hard gelatine capsules. All capsules were coated with a pH-responsive polymer, Eudragit S100 (Evonik Nutrition & Care GmbH, Darmstadt, Germany) designed to ensure targeted large intestinal delivery. The coating exhibits resistance to acidic environments, thereby maintaining capsule integrity during gastric transit. The methylacrylate polymer begins to dissolve at pH ≥ 6.5, facilitating the gradual release of the active ingredient upon arrival in the ileum and proximal colon, the intended site of delivery (32,33). The study capsules were manufactured, filled, and coated by Catalent Germany Schorndorf GmbH under GMP-compliant conditions.

### Measurements blood samples

Blood samples were collected using Becton Dickinson Vacutainer (K2E) tubes for plasma and Becton Dickinson Vacutainer (SST) tubes for serum (Belliver Industrial Estate. PL6 7BP, United Kingdom) and stored at −80°C. Safety profiling, including hematology, chemistry, glucose, bilirubin and hCRP, was conducted by standard clinical laboratory methods at Eurofins Biomnis. Riboflavin concentrations in plasma were measured via liquid chromatography, as described previously (32). Plasma sCD14 was determined using a quantitative sandwich enzyme-linked immunosorbent assay (CD14 ELISA Kit - Quantikine, R&D Systems Europe Ltd. Abingdon OX14 3NB, United Kingdom), in agreement with the manufacturer’s instruction. Plasma free thiol concentrations were measured using a plate-reader-based ultrasensitive fluorometric assay employing a Thiol Green Indicator, which produces a highly fluorescent adduct upon reaction with thiol groups (Free Thiol Assay Kit, Abcam plc, Cambridge United Kingdom). Quantification was based on a standard curve generated within the same assay plate. Pilot testing confirmed sample levels were within the assay’s dynamic range.

### Measurements fecal samples

#### Sample collection, DNA extraction and metagenome sequencing

After participants provided fecal samples to the laboratory, these were stored at −80°C and shipped on dry ice to BaseClear B.V. (Leiden, The Netherlands) for shotgun metagenomic sequencing. Genomic DNA extraction was performed using the ZymoBIOMICS™ 96 DNA Kit (Zymo Research, Irvine, CA, USA), and quantified using the Quant-iT™ dsDNA Broad Range Assay Kit (Thermo Fisher Scientific, Waltham, MA, USA). Paired-end shotgun sequencing was conducted on the Illumina NovaSeq 6000 platform. Quality control included adaptor trimming, and removal of PhiX reads using an in-house filtering protocol.

#### Taxonomic profiling of metagenome sequences

The raw sequence reads were processed through custom workflows to generate annotation count matrices. For taxonomic annotation, reads were filtered using Fastp (v0.20.1) based on quality, complexity, and length. Specifically, reads were discarded if they were shorter than 15 bp, contained more than 5 ambiguous bases (N), had more than 40% low-quality bases (Phred < Q15), or had sequence complexity below 30%. Taxonomic annotation was performed with Kraken2 (34) using a confidence threshold of 0.1, followed by species abundance estimation with Bracken (35), applying a minimum count threshold of 10 to reduce noise from low-abundance taxa. The Kraken2 database was built from the Unified Human Gastrointestinal Genome (36) version 2.01, release 202. Differential abundance of microbial taxa between treatment arms vs. placebo and across timepoints (baseline vs. week 4 and vs. week 12) was analyzed using ALDEx2 (ANOVA-Like Differential Expression). ALDEx2 utilized Monte Carlo sampling from the Dirichlet distribution to estimate technical variation and applied the Centred Log-Ratio (CLR) transformation to account for the compositional nature of the data. Microbiome alpha diversity was assessed using Shannon and evenness indices that quantify species richness and evenness, while beta diversity was evaluated through Jaccard and Bray-Curtis distance metrics to determine differences in overall microbial community composition and structure.

#### Computation of the Health-Associated Core Keystone (HACK) Index

The HACK index based on the top 17 taxa was calculated as recently described (37). Species-level taxonomic profiles were used and converted to relative abundances following standard preprocessing, including harmonization of species names. To align the original NCBI-based HACK species list with the GTDB-based taxonomy (version 202), a mapping between naming systems was created. In cases where multiple GTDB genome-level variants corresponded to a single NCBI species (e.g., *Faecalibacterium prausnitzii* mapped to *F. prausnitzii_C* and *F. prausnitzii_D*), their abundances were summed. Sixteen of the seventeen original HACK species were successfully matched while *Oscillibacter* sp. 57_20 could not be mapped and was therefore excluded from the index calculation. For each sample, the Hack index was determined by first assigning the sample to its subject and restricting the analysis to species consistently detected across all samples from that subject. Within this subset, species were ranked according to their relative abundance, and the HACK index was defined as the mean rank of the available top 17 HACK species.

#### Computation of the Dysbiosis Index

To assess global microbiome composition and stability, the dysbiosis index using the method developed by the EMBL Enterotype project (https://enterotype.embl.de/) was computed (38). This index quantifies the deviation of a sample’s genus-level microbial profile from a healthy reference centroid derived through enterotype-based clustering with higher values indicating greater compositional divergence from balanced enterotype structures. Species-level taxonomic profiles were used for computation as defined earlier and aggregated to the genus level. Genus names were formatted to match the required input conventions of the EMBL online tool, and only genera with valid mappings to the GTDB taxonomy were retained. Relative abundances were calculated per sample, and the resulting genus-level matrix was submitted to the EMBL Enterotype assignment webtool.

#### Computation of riboflavin prototrophic fraction

Individual species with available metabolic models from the highly curated AGORA2 (https://www.nature.com/articles/s41587-022-01628-0) and the large APOLLO resources (https://pmc.ncbi.nlm.nih.gov/articles/PMC10592896/) were analyzed using a Flux Balance Analysis (FBA) simulation to determine their ability to grow under unrestricted conditions with and without riboflavin, enabling classification of their dependency on external riboflavin supply. AGORA2 models were selected by GTDB name matches to species identified in this study. The APOLLO models were selected by calculating the best Average Nucleotide Identity (ANI) score against the top 100 most abundant species with ≥95% ANI and ≥75% alignment coverage. The prototrophic fraction was calculated as sum of relative abundance of prototrophs divided by sum of relative abundance of prototrophs and auxotrophs.

#### Microbial network analysis

Species-level taxonomic profiles were preprocessed by adding pseudo-counts, converting to relative abundances, filtering low-prevalence taxa, and applying the centered-log ratio (CLR) transformation. Microbial association networks were constructed for each treatment group (placebo, riboflavin at 1.4 mg, 10 mg, 75 mg) at week 0, 4, and 12 using SPIEC-EASI (39) via the NetCoMi R package version 1.2.0 (40), resulting in 12 networks. Associations were converted into signed dissimilarity values and then sparsified by applying an edge-weight threshold of >0.1. The keystone taxa for each network were prioritized based on their normalized degree centrality, closeness centrality and betweenness centrality, based on the work from Berry et al. (41). The cut-off was set at 40, corresponding to approximately the top 5% of the most influential nodes in the network. To decode the modular structure of the networks, the Louvain algorithm (42) was applied to identify the modules that maximize modularity within each constructed network.

#### Functional profiling of metagenome sequences

Raw metagenome sequence reads were filtered to remove human genomic contamination using Bowtie 2 v. 2.4.2 (43) against the GRCh38.p14 reference human genome. Reads were trimmed to remove adapters and to remove low-quality bases using Adapter Removal v. 2.3.1 (44). High-quality non-host (HQNH) read pairs (≥100 bp) were retained, yielding an average of 29.2 million reads per sample. Functional profiling was executed by using the CHAMP (45) pipeline including The Clinical Microbiomics Human Microbiome Reference HMR05 MAG-based with 6,567 prokaryotic and 244 eukaryotic species as a reference (Cmbio, Copenhagen, Denmark). The workflow followed the same methodology to calculate KEGG module abundances as described recently (46,47).

#### Short-Chain Fatty Acid (SCFA) analysis

Fecal samples were stored at −80 °C and shipped on dry ice to MS-Omics (Vedbæk, Denmark) for SCFA quantification. Sample preparation involved homogenization of 300 mg of fecal material with ultrapure water at a 4:1 weight-to-volume ratio, via bead beating (2 min at 30 Hz) followed by centrifugation (6,000 × g for 10 min at 4 °C) and filtration. Obtained supernatant were acidified and spiked with hydrochloric acid, and deuterium-labelled internal standards SCFA were quantified using a gas chromatograph (GC 7890B, Agilent Technologies, Santa Clara, CA, USA) equipped with a Zebron™ ZB-FFAP capillary column (30 m × 0.25 mm × 0.25 µm, Phenomenex, Torrance, CA, USA) coupled to an Agilent 5977B quadrupole mass spectrometer. Instrument control and data acquisition were performed with ChemStation software (Agilent Technologies). Raw data were exported in netCDF format and subsequently processed using PARADISe (MATLAB R2021b MathWorks Inc., Natick, MA, USA), as described previously (48). Total SCFA content was calculated as the sum of acetic acid, formic acid, propanoic acid, 2-methylpropanoic acid, butanoic acid, 3-methylbutanoic acid, pentanoic acid, 4-methylpentanoic acid, hexanoic acid, and heptanoic acid.

#### Calprotectin and pH analysis

Fecal samples were stored at 4□°C prior to shipment to Eurofins Biomnis for analysis. Fecal calprotectin concentrations were quantified using a fluoroenzyme immunoassay (FEIA, Thermo Fisher Scientific, Waltham, MA, USA) conducted on the Phadia™ 250 platform, employing the Phadia ELiA™ 2^nd^ Generation system. Fecal pH was assessed using a colorimetric strip-based method with Siemens test strips (Siemens Healthineers, Erlangen, Germany), followed by automated reading on a Siemens reader (COFRAC-accredited).

#### Food frequency, Gastrointestinal symptoms and Quality of life questionnaires

Food Frequency Questionnaires (FFQ) were used to assess the habitual dietary intake and to determine if the participant’s diet remained stable over time. Using nutrition analytical software Nutritics (Nutritics Ltd., Swords, Ireland) the daily fiber intake, as well as energy, carbohydrates, protein and fat were calculated. Gastrointestinal symptoms were assessed using the Gastrointestinal Symptom Rating Scale (GSRS, UK-E2, AstraZeneca AB, Södertälje, Sweden) (49). GSRS utilizes a 7-point rating scale, depending on the intensity and frequency of GI symptoms experienced during the previous weeks. A higher score indicates more inconvenient symptoms. The Short-Form Health Survey (SF-36v2, 4-week recall version, QualityMetric Inc., RI, USA, License QM055304) was used to assess the quality of life (50). It covers 8 domains; physical functioning, limitations due to physical health, bodily pain, general health, vitality, social functioning, and limitations related to emotional and mental health. The Physical Component Score (PCS) was derived from physical functioning, limitations due to physical health, bodily pain and general health perceptions while Mental Component Score (MCS) was derived from mental health, limitations due to emotional problems, social functioning and vitality. Domain scores were calculated by recoding each item to a 0–100 scale and averaging across items within each domain.

### Sample size calculation and statistical analysis

#### Sample size

The anticipated effect size and standard deviation were based on data from a previous study that assessed the impact of 75□mg riboflavin over four weeks (32). Using these findings, a sample size of 79 participants per group was determined to be necessary to detect a 1.2% difference in the relative abundance of selected bacterial species between each riboflavin dose group and placebo, assuming a shared standard deviation of 2.3%, 80% statistical power, and a significance level (α) of 0.017. This alpha level reflects a Bonferroni adjustment, dividing the conventional 0.05 threshold across the three dose comparisons to placebo to ensure a conservative and robust estimate for sample size determination. To accommodate a projected 10% dropout rate, the target enrolment was increased to 87 participants per group, resulting in a total sample size of 348 individuals. Missing, unused, or anomalous data were addressed using the Last Observation Carried Forward (LOCF) approach within analyses of the Intention-to-Treat (ITT) population (Supplementary Figure S1). Since the expected dropout rate was below 10%, no participant replacement strategy was implemented during the course of the study.

#### Statistical analysis

Efficacy analysis assessed (1) whether there were significant within-group changes from baseline to the end of the intervention in either the riboflavin groups or the placebo group, and (2) whether there were significant between-group differences (i.e., whether changes over time in a riboflavin group differed from those observed in the placebo group).

For relative taxa abundance and microbial alpha diversity, paired *t*-tests (or Wilcoxon signed rank tests) were used while Permutational Multivariate Analysis of Variance (PERMANOVA) was conducted to test for significant differences in community composition between groups (beta diversity). Resulted *p*-values were not adjusted for multiple testing. For the HACK Index, Dysbiosis Index, and riboflavin prototrophic fraction, between-group differences were analyzed using analysis of covariance (ANCOVA) with baseline values as covariates to evaluate the statistical significance of differences between the placebo and riboflavin groups while within group changes were assessed using the Wilcoxon signed-rank test. Resulted *p*-values were adjusted for multiple testing by Bonferroni correction. Between group comparisons of network topological properties, including node count, edge density, clustering coefficient, modularity, positive edge percentage, and natural connectivity were performed using one-tailed *t*-tests. Keystone taxa were identified based on centrality metrics (41) while modular structures were detected using the Louvain algorithm (42). Statistical analysis of KEGG module abundances was performed using LinDA (51), using a linear regression framework with compositional bias correction. Abundances were log-transformed, with zeros replaced by pseudo-counts. Bias-corrected coefficients were interpreted as log2 fold changes. Modules present in <10% of samples were excluded. For all other non-microbiome related endpoint variables measured at week 4 and 12, paired *t*-tests (or Wilcoxon signed rank tests) were used to test for within-group changes while ANCOVA, with baseline values included as covariates were used for between group changes. Planned comparisons were conducted between each active treatment group and the placebo group. In instances where assumptions of normality were violated, Quade’s rank ANCOVA, a non-parametric alternative was employed.

## Results

### Study population

Numbers of participants at each stage of the trial are shown in Supplementary Figure S1. Analysis of the baseline microbiome profiles at week 0 across the four groups, using both alpha and beta diversity metrics, showed no significant differences, indicating that the microbiome composition was comparable among groups and supporting the validity of randomization. In addition, no statistically significant differences were observed in reported energy (kcal), carbohydrate (g), protein (g), or fat (g) intake between groups from baseline to week 12.

### Primary endpoint - relative taxa abundance

Between-group analysis revealed several significant changes in relative taxa abundance after 12 weeks in all riboflavin groups compared to placebo, with the most pronounced effects observed in the 10 mg riboflavin group. In addition, several earlier changes were already detectable at week 4 when compared with placebo (P≤0.05, effect size ≥0.15, Fig. 1, Supplementary Table S2).

**Fig. 1.**
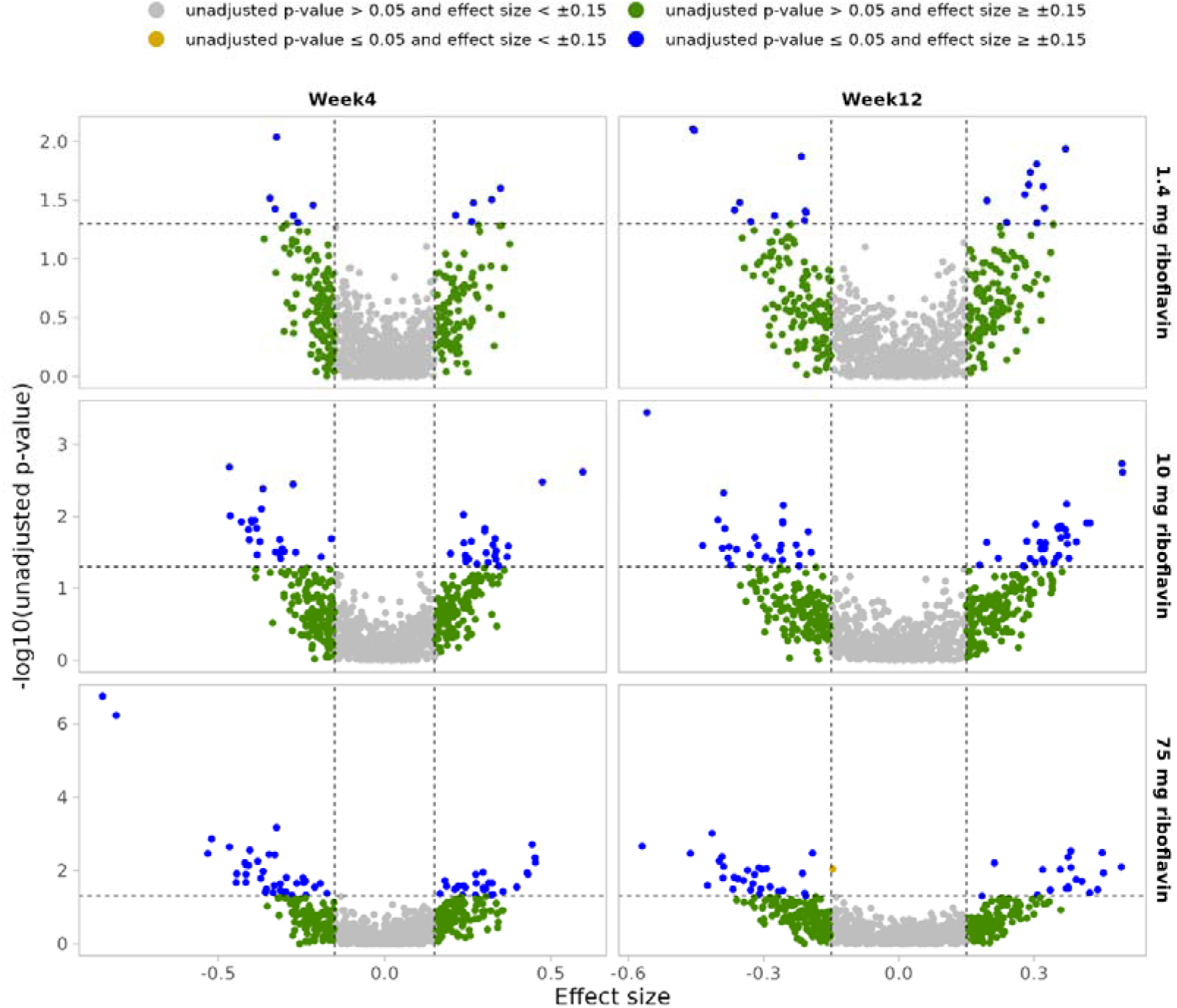
Effects of colon-delivered riboflavin on fecal microbial composition between groups. Volcano plots illustrate the between-group differential abundant species at week 4 and week 12 when compared to placebo. Grey dots: unadjusted *p-*value > 0.05 and effect size < +/- 0.15; Orange dots: unadjusted *p-*value ≤ 0.05 and effect size < +/- 0.15; Green dots: unadjusted *p-*value > 0.05 and effect size ≥ +/- 0.15; Blue dots: unadjusted *p-*value ≤ 0.05 and effect size ≥ +/- 0.15.

Increases in relative species abundance across all six comparisons clustered most consistently within the phylum *Bacillota* (formerly *Firmicutes*), particularly within class *Clostridia,* especially the orders *Lachnospirales* (family *Lachnospiraceae*) and *Oscillospirales* (families *Acutalibacteraceae, Ruminococcaceae, Oscillospiraceae*) (Fig. 1, Supplementary Table S2). Recurrent increase in *Lachnospiraceae* taxa include the genera *Blautia_A, Faecalimonas*, or *Mediterraneibacter,* alongside *Oscillospirales* lineages such as *Ruminococcus_E.* These genera appeared in multiple dose–time combinations, suggesting a shared, dose-independent pattern toward enrichment of acetate and/or butyrate-associated *Bacillota*. Also the phylum *Bacteroidota* (notably the family *Bacteroidaceae*) became evident most clearly at 10 and 75 mg at week 12, with increases in the genera *Bacteroides*, *Phocaeicola*, and *Prevotella.* By contrast, *Pseudomonadota* increases were largely confined to the 75 mg groups (e.g., *Klebsiella variicola* at Week 4; *Escherichia* spp. and *Raoultella* at Week 12), while also *Verrucomicrobiota* were observed only at 75 mg Week 12.

When examining individual doses at different time points *vs.* placebo, the increases in relative abundance for 1.4 mg of riboflavin at week 4 were relatively concentrated within *Lachnospiraceae*/ *Acutalibacteraceae* (e.g., *Anaerostipes*, *Eubacterium_F*, *Pseudoruminococcus*), whereas by week 12 the increases broadened to also include *Lachnospira*, *Anaerobutyricum hallii*, and *Ruminococcus_B gnavus*. At 10 mg of riboflavin, there was a rich *Lachnospiraceae* response (multiple *Blautia_A, Roseburia, Lachnospira, Mediterraneibacter*) and some effects on Negativicutes (*Acidaminococcus*, *Succiniclasticum*) and *Prevotella* spp. at week 4, while by week 12, there was the widest increase in *Bacteroidaceae* (e.g., *Bacteroides stercoris/acidifaciens/togonis*, *Phocaeicola dorei/ massiliensis/sartorii*) alongside increases in *Anaerobutyricum* (*hallii*, *soehngenii*) and recurring *Ruminococcus_E/Negativibacillus*. With 75 mg at week 4, there were increases in *Alistipes, Christensenellales* and *Ruminococcus_E bromii*, while at week 12, the most pronounced increases included elevated levels of *Lachnospiraceae/Oscillospirales* with *Bacteroidaceae, Pseudomonadota* (*Escherichia*, *Raoultella*), *Lactobacillales* (*Lacticaseibacillus rhamnosus*), and a *Verrucomicrobiota* signal.

Significant decreases in relative species abundance across all riboflavin arms *vs.* placebo, clustered in the phyla *Bacillota_A (Firmicutes_A,* class *Clostridia*) and *Actinobacteriota* (class *Coriobacteriia*). Within *Clostridia,* decreases were observed in the order *Oscillospirales*, spanning *Acutalibacteraceae* (e.g., UBA1417, UMGS1487, UMGS1591), *Oscillospiraceae* (CAG-83, CAG-103, NK3B98, UBA5446, CAG-110) and *Ruminococcaceae* (including multiple *Faecalibacterium species* at 75 mg), but also extended to the *Christensenellales* order at 10 and 75 mg. Additional *Bacillota* reductions were observed for *Anaerotignum, Flavonifractor, Clostridium_Q* symbiosum, and *Intestinimonas*. These reductions in relative abundance across all groups were accompanied by consistent decreases in *Coriobacteriia*, more specifically *Eggerthellaceae*, including recurring decreases in genera like *Adlercreutzia, Slackia*, *Collinsella, Senegalimassilia*, and provisional lineages (CAG-1427, CAAEEV01, UMGS124), as well as *Olsenella* at 75 mg, week 12. Signals beyond this overall clustering were more dose–time specific (Fig. 1, Supplementary Table S2).

Within-group analysis confirmed that the most pronounced effects were observed in the 10 mg riboflavin group at week 12 with the highest number of species significantly changed over time (Fig. 2, Supplementary Figure S2 and Supplementary Table S3). Across all intervention arms, similar changes in relative abundances were observed with increases in the phylum *Bacillota_A* (*Clostridia*, *Lachnospiraceae*), notably the genera *Blautia_A, Dorea, Lachnospira, Coprococcus, Roseburia*, *Anaerostipes*, together with increases in some *Ruminococcaceae* and genera belonging to the phylum *Bacteroidota,* family *Bacteroidaceae* (*Phocaeicola, Bacteroides*, *Prevotella*). Within-group analyses of decreased taxa showed time- and dose-dependent downshifts in addition to small placebo drifts. In placebo, week 0 to 4 modest declines were observed in *Oscillospiraceae* (CAG-83, CAG-103), *Ruminococcaceae* (*Faecalibacterium*), and *Collinsella,* while at week 12 *Ruthenibacterium lactatiformans* and *Christensenellales* were also reduced. Against this background, riboflavin arms exhibited coherent reductions concentrated in Bacillota, class *Clostridia*, notably *Lachnospirales* (*Lachnospiraceae*) and *Oscillospirales (Ruminococcaceae, Oscillospiraceae*), together with prominent *Actinobacteriota (Coriobacteriia*) signals (Fig. 2, Supplementary Figure S2 and Supplementary Table S3).

**Fig. 2.**
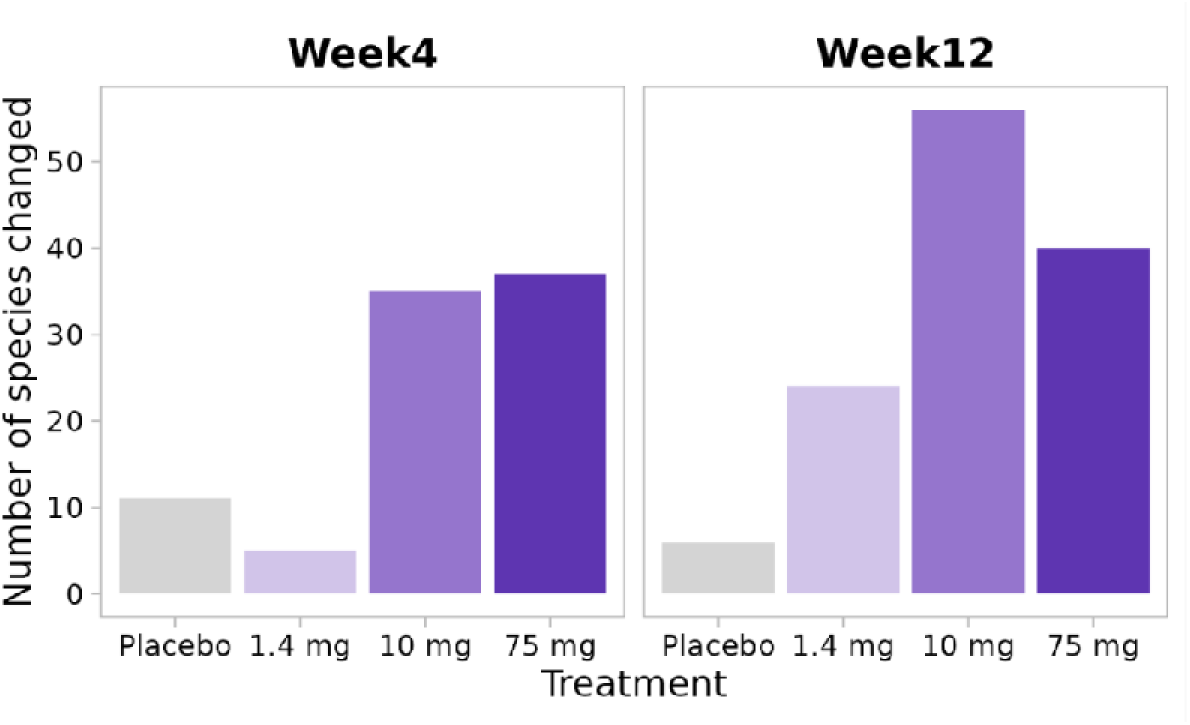
Effects of colon-delivered riboflavin on number of species changed within groups. Bar charts represent the number of species that significantly changed (increase or decrease) in relative abundance over time within each group at week 4 and week 12 compared to baseline.

### Secondary endpoints

#### Microbial beta and alpha diversity

There were no significant differences between or within groups in beta diversity. However, analysis of fecal microbial alpha diversity revealed a significant within-group difference with an increase in evenness and the Shannon index for riboflavin at 10 mg at week 12 when compared to baseline (Fig. 3A and B).

**Fig. 3.**
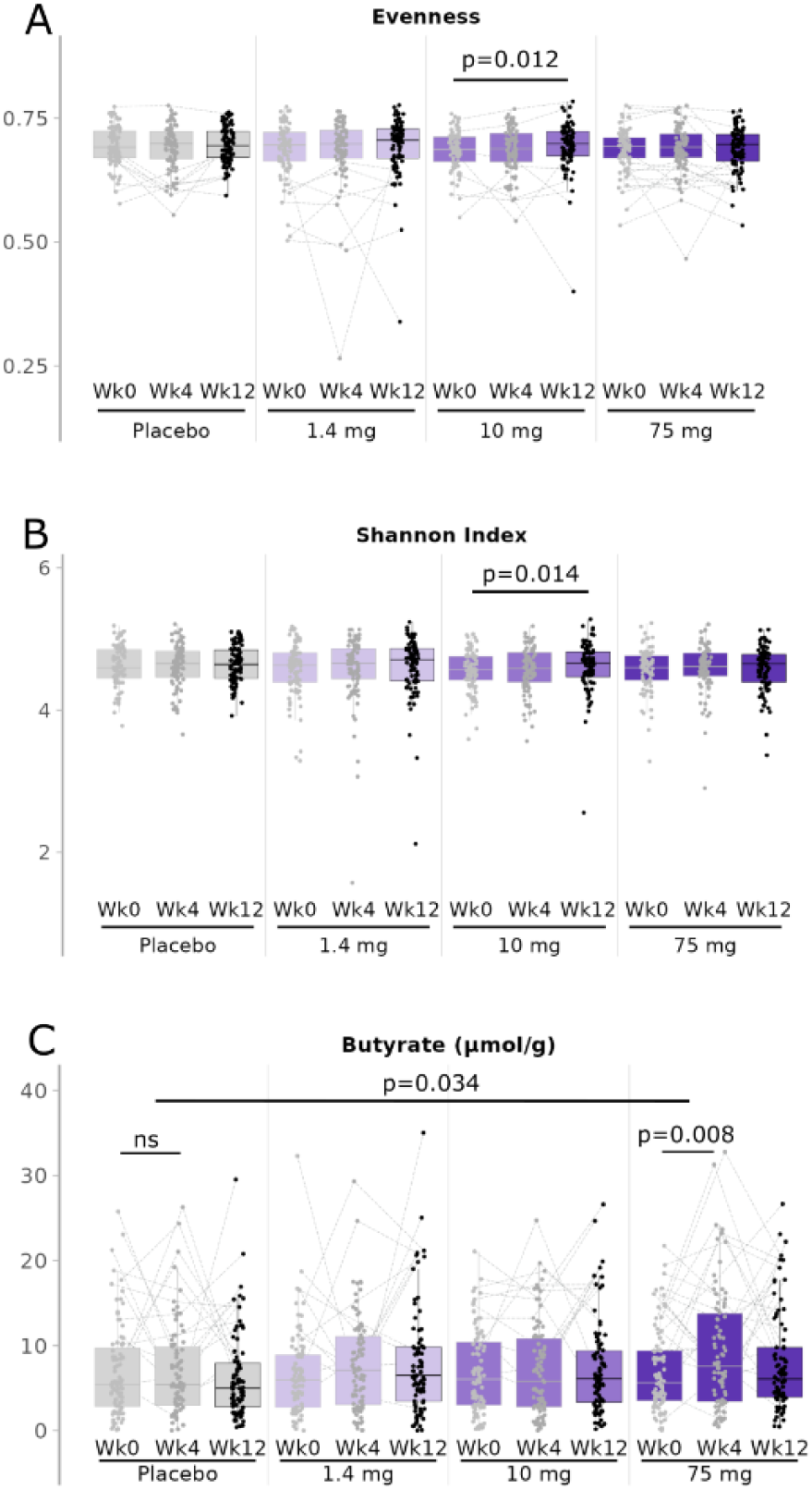
Effects of colon delivered riboflavin on fecal microbial alpha diversity and fecal butyrate. Data represent box-and-whisker plots of microbial evenness (A), Shannon index (B) and fecal butyrate concentration (C). Box plots show median (center line), interquartile range (box), and whiskers extending to 1.5x the interquartile range. Points beyond the whiskers represent outliers. ns, not significant, *p-*value ≤ *0.05*, significant changes.

#### *Fecal* short-chain fatty acid (SCFA)

Riboflavin at 75 mg significantly increased fecal butyrate concentrations at week 4 within groups when compared to baseline (*p = 0.008*) as well as between groups when compared to placebo (*p = 0.034,* Fig. 3C). No other significant differences in fecal SCFA (propionate, acetate, total SCFAs) were observed either between or within-groups for any riboflavin doses.

#### Fecal pH

There were no significant between- or within-group differences in fecal pH at week 12 for any of the riboflavin doses (Table 1).

**Table 1.** Changes in blood and fecal biomarkers as well as questionnaires in response to colon-delivered riboflavin at 1.4, 10 and 75 mg/day.

| Parameters | placebo |  | 1.4 mg |  | 10 mg |  | 75 mg |  |
| --- | --- | --- | --- | --- | --- | --- | --- | --- |
|  | Wk 0 | Wk 12 | Wk 0 | Wk 12 | Wk 0 | Wk 12 | Wk 0 | Wk 12 |
| Faecal pH | 6.97 (0.07) | 6.98 (0.06) | 6.95 (0.06) | 6.87 (0.07) | 7.04 (0.07) | 7.00 (0.07) | 6.87 (0.06) | 6.90 (0.06) |
| Faecal calprotectin<br>in µg/g | 15.72 (3.88) | 26.78 (7.58) | 17.08 (3.59) | 18.12 (3.80) | 9.74 (1.38) | **22.30 (4.14)\*** | 17.94 (4.25) | **30.71 (5.94)\*** |
| Plasma hs-CRP<br>in mg/L | 1.54 (0.22) | 1.52 (0.21) | 1.79 (0.34) | 2.17 (0.77) | 1.26 (0.14) | 1.54 (0.25) | 1.32 (0.14) | 1.87 (0.26) |
| Plasma Free Thiol<br>in nM | 101.40 (3.82) | 115.48 (3.50) | 100.41 (3.88) | **118.77 (3.00)\*** | 101.20 (3.72) | 112.90 (2.78) | 104.80 (3.97) | **115.84 (3.08)\*** |
| Plasma sCD14<br>in pg/ml | 1568.07 (52.49) | **1197.52 (45.75)\*** | 1734.64 (74.55) | **1278.54 (44.85)\*** | 1682.16 (63.74) | **1262.22 (43.20)\*** | 1665.38 (65.61) | **1774.44 (473.12)\*** |
| Plasma riboflavin<br>in ng/mL | 6.65 (0.78) | 6.65 (0.62) | 7.61 (1.19) | 8.35 (1.29) | 6.17 (0.49) | 6.37 (0.40) | 6.80 (0.84) | **7.28 (0.68)\*#** |
| GSRS total score | 1.23 (0.03) | 1.27 (0.03) | 1.21 (0.03) | 1.23 (0.03) | 1.26 (0.03) | 1.32 (0.03) | 1.23 (0.03) | 1.29 (0.03) |
| Quality of Life –<br>mental component summary | 56.02 (0.48) | 55.96 (0.48) | 55.32 (0.56) | 55.43 (0.60) | 55.62 (0.49) | 55.79 (0.49) | 55.55 (0.52) | 55.51 (0.53) |
| Quality of Life –<br>physical component summary | 56.22 (0.39) | 56.53 (0.43) | 56.20 (0.52) | 55.96 (0.52) | 56.78 (0.47) | 56.28 (0.52) | 56.65 (0.53) | 56.05 (0.46) |
| Stool consistency | 3.61 (0.10) | 3.61 (0.10) | 3.52 (0.10) | 3.74 (0.09) | 3.57 (0.10) | 3.47 (0.10) | 3.74 (0.09) | 3.74 (0.10) |
| Bowel Movements | 1.17 (0.02) | 1.20 (0.03) | 1.17 (0.03) | 1.17 (0.03) | 1.14 (0.02) | 1.21 (0.04) | 1.21 (0.03) | 1.23 (0.03) |

Data are shown as mean (SD)
\* indicate significant difference within groups $p < 0.05$ while # indicate significant difference between groups versus placebo ( $p < 0.05$ ).

#### Fecal calprotectin

There were no significant between-group differences in fecal calprotectin for any of the riboflavin doses. A significant within-group increase, within the normal clinical ranges, was observed at week 12 when compared to baseline for riboflavin at 10 and 75 mg (*p* ≤ *0.05 respectively,* Table 1).

#### Plasma high-sensitivity C-reactive protein (hs-CRP)

No significant differences in hs-CRP concentrations were observed either between or within-groups at week 12 for any riboflavin doses (Table 1).

#### Plasma free thiols

There were no significant between-group differences in plasma free thiols for any of the riboflavin doses. However, we observed a significant within-group difference with an increase in plasma free thiols at week 12 when compared to baseline for riboflavin at 1.4 and 75 mg (*p* ≤ *0.05 respectively,* Table 1).

#### Plasma sCD14

There were no significant between-group differences in sCD14 at week 12 for any of the riboflavin doses. However, we observed significant within-group changes with decreased sCD14 concentrations at week 12 when compared to baseline for the placebo group as well as the groups receiving riboflavin at 1.4 and 10 mg. In contrast there was a slight but significant increase in sCD14 with riboflavin at 75 mg (*p* ≤ *0.05* for all, Table 1).

#### Plasma riboflavin concentrations

Riboflavin at 75 mg significantly increased plasma riboflavin concentrations at week 12 within groups when compared to baseline as well as between groups when compared to placebo (*p* ≤ *0.05*, Table 1).

#### Gastrointestinal Symptoms Rating Scale (GSRS), Quality of life short form Health Survey with 36 questions (SF-36) and stool consistency and frequency

No significant between or within-group differences in the GSRS total score, the SF-36 physical health and mental health component score and stool consistency or frequency were observed at week 12 for any riboflavin dose (Table 1).

### Exploratory Endpoints

#### HACK index and Dysbiosis index

The HACK index, which assigns a score for each species reflecting association with 17 selected keystone taxa strongly linked to a beneficial, resilient microbial community, was not significantly different between groups at week 4 or week 12 for any riboflavin doses. However, a significant within-group difference with an increase for riboflavin at 10 mg was observed at week 12 when compared to baseline (*p = 0.045*, Fig. 4A). There was also no between group difference for the dysbiosis index, which quantifies the deviation of a sample’s genus-level microbial profile from a healthy reference centroid derived through enterotype-based clustering. However, we observed a significant within-group change with a decrease for riboflavin at 1.4 mg at week 12 when compared to baseline (*p = 0.007*, Fig. 4B).

**Fig. 4.**
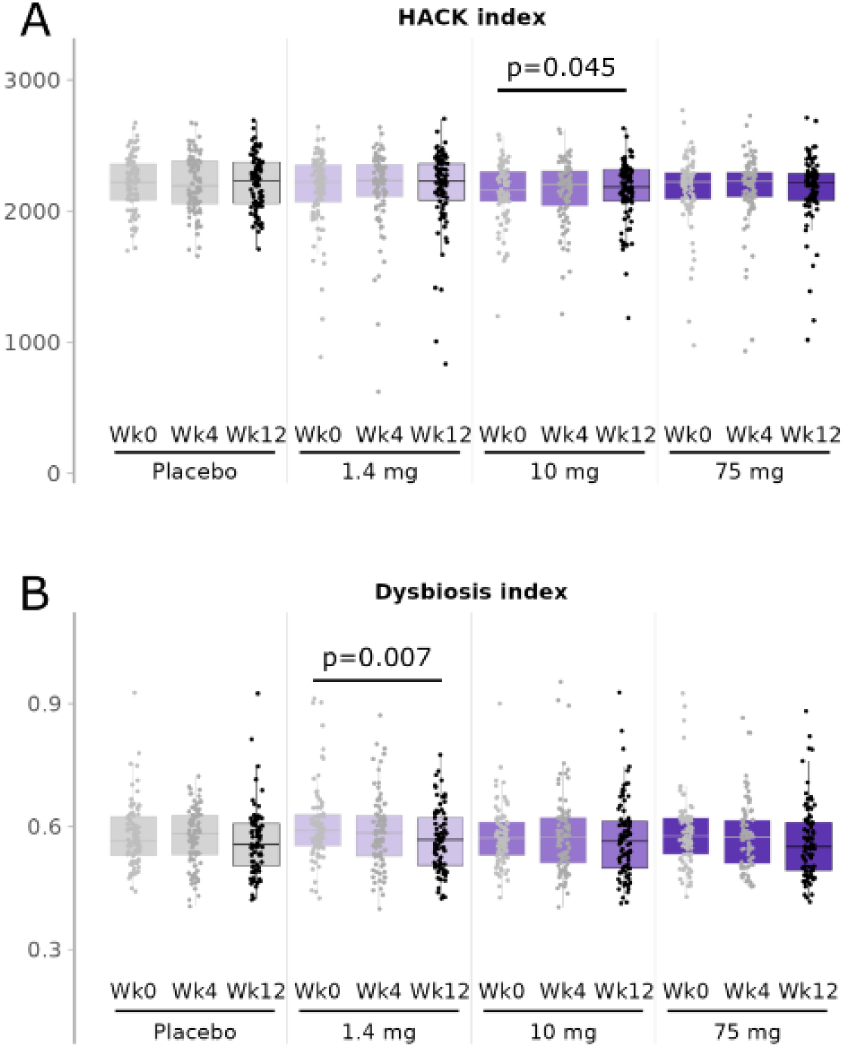
Effects of colon delivered riboflavin on microbiome health indices. Data represent box-and-whisker plots of the Hack index (A) and Dysbiosis index (B). Box plots show median (center line), interquartile range (box), and whiskers extending to 1.5x the interquartile range. Points beyond the whiskers represent outliers. *p-value* ≤ *0.05*, significant changes.

#### Modular network structure and keystone taxa

When comparing modular network structure and performance across groups per time point, a modest but statistically significant difference was observed with riboflavin at 1.4 mg at week 4 when compared to placebo with an increase in edge density and natural connectivity (*p* ≤ *0.05*, Supplementary Table S4).

Keystone taxa patterns were investigated by computing modular network structures highlighting keystone taxa prioritized according to their centrality scores for each dose and timepoint. Temporal analysis within intervention groups showed limited stability of keystone taxa in the placebo group, with only eight taxa consistently detected across all timepoints (Fig 5A). The number of unique keystone taxa in the placebo group also declined from 20 at baseline to 17 at week 12. In contrast, the 1.4 mg colon-delivered riboflavin group maintained 11 shared keystone taxa across timepoints, accompanied by a gradual increase in unique keystone taxa. The 10 mg group retained nine core keystone taxa and reached 20 unique keystone taxa by week 12. Most prominently, the 75 mg group exhibited the highest persistence, with 13 keystone taxa consistently present across timepoints, along with the largest rise in unique keystone taxa, increasing from 8 at baseline to 17 at week 12 (Fig. 5A). Figure 5 B illustrates a modular network structure observed after 12 weeks of supplementation with 1.4 mg riboflavin. Notably, several identified keystone species also showed increased relative abundance within or between groups, including *Lachnospiraceae* genera such as *Blautia_A* and *Dorea* (Fig. 5A, Supplementary Table S2 and 3).

**Fig. 5.**
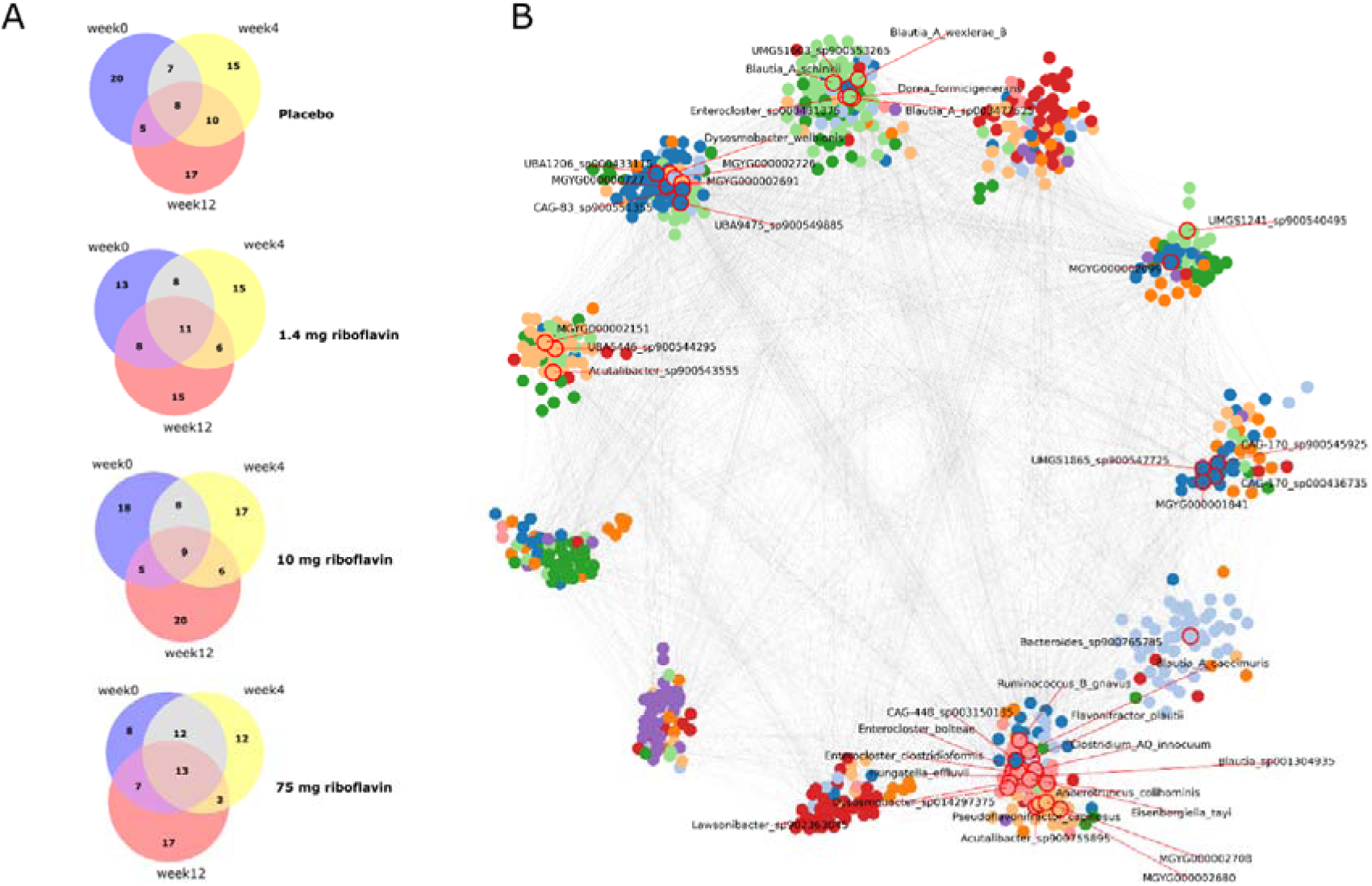
Effects of colon delivered riboflavin on keystone species distribution and modular network structure. Data show a Venn diagram of overlapping and unique keystone taxa within each treatment group over time (A) and an example of a modular network structure highlighting keystone taxa prioritized based on their centrality scores within each network in response to riboflavin at 1.4 mg at 12 week (B).

#### Functional KEGG modules and riboflavin prototrophic/auxotrophic fraction

No significant differences in KEGG module abundance were detected between groups. However, within-group analysis revealed significant changes at week 12 in 6, 86, and 124 KEGG modules at 1.4 mg, 10 mg, and 75 mg of riboflavin supplementation, respectively while no changes were observed in the placebo group. Among the significantly altered modules, the majority were upregulated with 100% (6/6) at 1.4 mg, 59% (51/86) at 10 mg, and 77% (96/124) at 75 mg (Fig. 6). Similar patterns were also evident as early as week 4. When filtering to only include modules that showed a false discovery rate (FDR) < 0.01 in at least one comparison, a total of 36 KEGG modules emerged as being most distinct. Notably, among this subset a module involved in riboflavin biosynthesis was included with 10 mg of riboflavin supplementation (Fig. 7A,C).

**Fig. 6.**
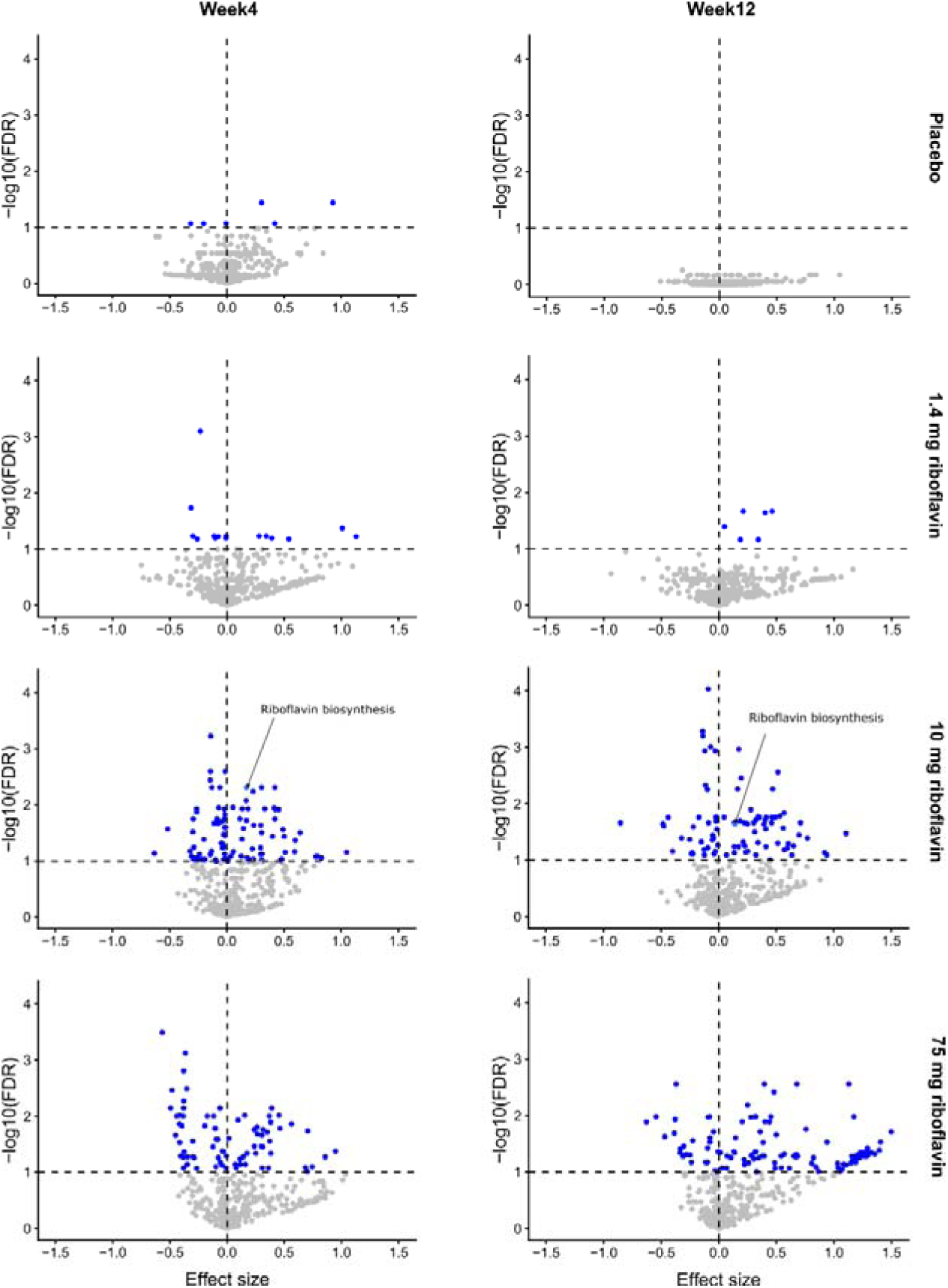
Effects of colon-delivered riboflavin on fecal microbial metabolic capability. Volcano plots summarize the within-group differential abundant KEGG modules at week 4 and week 12 when compared to baseline. Each dot refers to one test for a module and is colored blue if significantly (FDR < 0.1) different, either enriched in baseline (effect size < 0) or enriched at endpoint (effect size >0). Non-significant modules are indicated by grey dots. The riboflavin biosynthesis module (MF00125) is highlighted if significant. The dotted horizontal line indicates an FDR cutoff of 0.1.

**Fig. 7.**
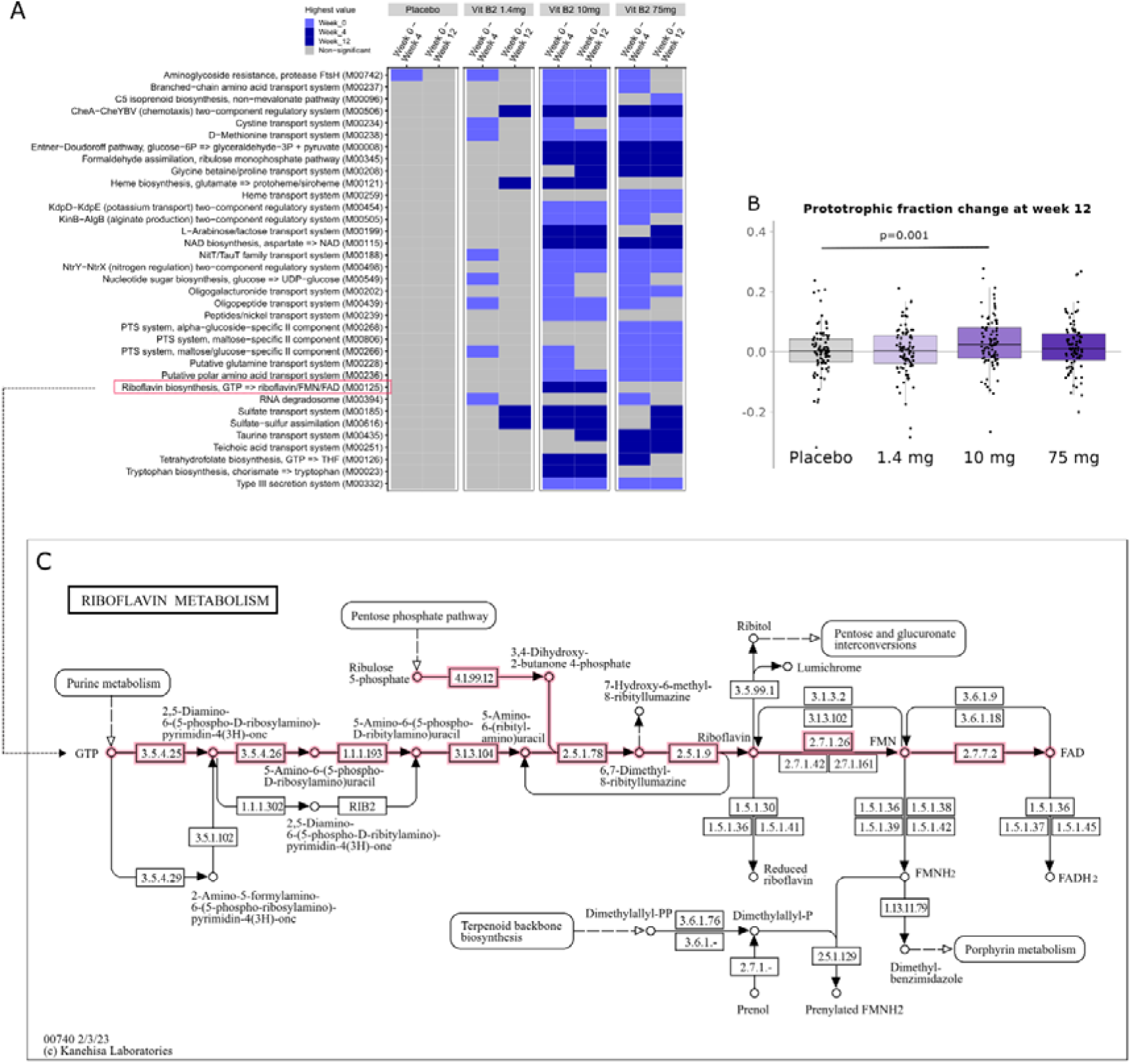
Effects of colon-delivered riboflavin on fecal microbial metabolic capability and riboflavin prototrophic fraction. Data represent a binary heat map showing changes in 34 KEGG modules with an FDR <0.1 in at least one comparison (A), changes in the fractions of riboflavin prototrophic species (B), and the riboflavin metabolism pathway (KEGG map00740, © Kanehisa Laboratories) (46) with the riboflavin biosynthesis module M00125 highlighted in light red (C). Light blue colored modules in (A) indicate highest value at week 0 (i.e. downregulated during intervention); Dark blue colored modules indicate highest value at week 4 or 12. (i.e. upregulated during the intervention). Grey colored modules are not significant.

Furthermore, when analyzing individual species with available metabolic models to determine their ability to grow with and without riboflavin, 267 species representing 76.5% of total abundance were identified by name matching to AGORA2 (176) and genome-based matching to APOLLO (91). The models of these species demonstrated *in silico* growth under unrestricted conditions, and for 245 species their trophy status could be unambiguously determined, of which 86 species could grow *in silico* when riboflavin was removed. Hence, 86 species are considered riboflavin prototroph and 159 species riboflavin auxotroph (Supplementary Table S5). When investigating the effect of colon delivered riboflavin on the balance of the prototrophs vs. auxotroph species fractions, a significant increase in the prototrophic fraction was found with the 10 mg colon delivered riboflavin (*p = 0.001*), whereas no increase was observed with placebo, 1.4 mg or 75 mg of colon delivered riboflavin (Fig. 7B).

### Safety endpoints

Over the 12-week period, 4 subjects discontinued the study due to consent withdrawal, adverse events (AEs), or illness. A total of 148 participants (42.5%) experienced at least one AE, with 22 AEs (6.3%) deemed mild grade and considered related to the intervention. Among those, the most frequently reported AE was gastrointestinal discomfort, which occurred evenly across all intervention groups, including placebo. Regarding fecal calprotectin, 17 AEs (defined as fecal calprotectin >100 µg) were recorded after the study including 5 in the placebo group, 2 in the 1.4 mg riboflavin group, 5 in the 10 mg riboflavin group, and 5 in the 75 mg riboflavin group (Supplementary Figure S3). None were deemed related to the product, and as noted earlier, the average calprotectin levels across all groups remained within the normal clinical range both before and after the intervention. Laboratory evaluations, including hematology and biochemistry parameters, also showed no clinically significant changes and remained within normal reference ranges. Similarly, vital signs and physical examination findings showed no meaningful deviations over time, thus, overall, the IP was considered safe and well tolerated.

## Discussion

The present study shows that colon-delivered riboflavin at 1.4 mg, 10 mg, and 75 mg modulates gut microbiome composition, metabolic activity, ecosystem performance and network structure in a dose-specific manner in healthy older adults. Although no significant changes were observed between groups in overall community-wide diversity metrics (alpha and beta diversity), several dose-specific effects emerged from a detailed within and between group comparisons and exploratory endpoint analyses: 1) Colon-delivered riboflavin at all doses significantly altered the relative abundance of several microbial taxa when compared with placebo. 2) The most pronounced effects on microbiome composition, function, and network structure were observed with the 10 mg dose at week 12, reflected by within group increases in alpha diversity, the largest rise in total species counts, elevated HACK index values indicating greater community resilience, and distinct shifts in KEGG module abundance, including enhanced riboflavin biosynthesis. 3) Supplementation with 75 mg riboflavin led to a transient increase in fecal butyrate concentrations at week 4 versus placebo. 4) The lowest dose (1.4 mg) significantly reduced the dysbiosis index within groups and modestly improved network structure across groups, while all three doses (1.4, 10, and 75 mg) influenced keystone species abundance. Collectively, these findings point to optimal dose of colon-delivered riboflavin at around 10 mg with additional dose specific effects at 1.4 mg and 75 mg.

Our clinical trial builds on a previous pilot study in 12 healthy adults, where 4 weeks of colon-delivered riboflavin at 75 mg/day altered gut microbiome composition (32). In that pilot investigation, we reported within-group increases in alpha diversity (observed number of species), a finding which is consistent with the within-group increases in evenness and Shannon index observed in the present trial. It also aligns with a recent analysis of gut microbial vitamin production pathways in relation to clinical and lifestyle factors using the deeply phenotyped Dutch Lifelines-DEEP cohort (N = 1,135) which found a strong positive association between the Shannon alpha diversity index and the riboflavin biosynthesis pathway (52). A more diverse microbiome is generally considered more resilient to perturbations such as antibiotics, pathogens, or dietary shifts. Greater taxonomic richness expands enzymatic capacity, supporting functional redundancy and metabolic flexibility. Higher alpha diversity has been commonly linked to healthier metabolic, immune, and inflammatory profiles, while reduced diversity is often associated with IBD, obesity, type 2 diabetes, and allergies (1,4,53–57).

Another consistent finding across the previous pilot study, the current trial, and a recent TIM-2 *in-vitro* study validating colonic delivery of riboflavin via the proprietary food-grade multi-unit particle system Humiome B2® (31,32), was the enrichment of acetate- and butyrate-producing *Clostridia*, including several taxa within the orders *Lachnospirales, Oscillospirales and Clostridiales (*family *Clostridiaceae).* Altogether, these findings indicate that colon-targeted riboflavin may stimulate taxa contributing to acetate and butyrate production. The most consistent change was an increase in *Blautia* (notably with 10 mg riboflavin at week 12), a genus whose species produce acetate that supports colonocyte function and serves as a substrate for butyrate cross-feeding. Higher *Blautia* counts are frequently linked to more favorable gut-microbiome composition and metabolic status (58–61).

A recurring pattern across studies was also an increase in *Coprococcus,* observed in the pilot trial (riboflavin alone arm and when combined with vitamin C) (32) and again in the current study at 10 mg of riboflavin at week 4 and week 12 relative to baseline. In addition, this signal was seen also in the *in vitro* study using the TIM-2 model (31). *Coprococcus* is notable for its depletion in depression and positive association with quality of life (62,63). An additional noteworthy finding, albeit observed for the first time only in the present trial, was an increase in the closely related *Anaerobutyricum hallii* and *A. soehngenii,* at both 1.4 mg and 10 mg by week 12, evident within groups and relative to placebo. Following recently completed, as well as ongoing, *in-vitro* and clinical studies, these strictly anaerobic genera, in particular *A. soehngenii,* are considered promising next-generation probiotic candidate due to their efficient lactate utilization, strong butyrate-producing capacity, and demonstrated benefits on metabolic health, including biomarkers relevant to cardiovascular function (64–68).

At the 75 mg riboflavin dose, we also noted an increase in *Alistipes* at week 4 compared to placebo, consistent with findings observed in our earlier pilot study at this dosage (32). *Alistipes* species are common gut commensals that can ferment amino acids and produce SCFAs such as acetate and propionate (69). As a relatively newly described genus of bacteria, its effect on human health seem not yet fully understood and context as well as strain dependent with contradictory evidence regarding its protective or harmful role for liver, cardiovascular or digestive health or the gut-brain axis (69). Another notable finding at the 75 mg riboflavin dose is the significant reduction in *F. prausnitzii* at both week 4 and week 12, which is consistent with findings from our earlier pilot study at the same dosage (32). However, Liu et al.(60) reported no major shifts in gut microbiota composition, including *F. prausnitzii*, following oral riboflavin supplementation (50–100 mg/day for 2 weeks). Similary, Martels et al. (70) found no increase in *F. prausnitzii* in Crohn’s disease patients receiving 100 mg riboflavin daily for three weeks. Together these human data contrast with results from previously reported *in-vitro* work by Harmsen and colleagues (17,18) showing that *F. prausnitzii* uses extracellular riboflavin as an electron shuttle to reduce oxygen and local lower redox potentials to survive in moderately oxygenated niches (e.g., near the mucosal surface). This mechanism is known as flavin-based extracellular electron transfer (FLEET) (19), and suggests that riboflavin may act as a selective growth factor for *F. prausnitzii*. Accordingly, a pilot open-label study in healthy volunteers found that oral supplementation with 100 mg riboflavin for 14 days increased fecal *F. prausnitzii* counts (71). Several factors may explain the discrepancies observed in the different studies: 1) *F. prausnitzii* is a species complex, with genetically and functionally distinct strains that vary in oxygen tolerance and riboflavin dependency (72). In fact, our metabolic modeling indicated that strains showing significant decreases, such as *F. prausnitzii_D* were riboflavin prototrophs (see Supplementary Table 2 and 3). 2) The FLEET mechanism may be detectable only *in-vitro* within monocultures or small microbial consortia, whereas *in-vivo*, within complex microbial communities, riboflavin-auxotrophic species with higher riboflavin affinity could outcompete *F. prausnitzii*. 3) The FLEET mechanism may act in a dose-dependent manner *in-vivo*, being effective at specific dosages, whereas higher doses appear to exert growth-inhibitory effects on *F. prausnitzii*.

The decline in *F. prausnitzii* also suggests that the week 4 increase in fecal butyrate with 75 mg riboflavin is more likely attributable to the observed broad stimulation of acetate- and/or butyrate-producing *Clostridia* rather than *F. prausnitzii*. A similar rise in fecal butyrate was reported also by Liu *et al.* (26), who noted elevated butyrate after two weeks of oral riboflavin supplementation at 100 mg/day, along with increased acetate at 50 mg/day. These effects occurred without major shifts in overall gut microbiota composition, including *F. prausnitzii*. However, the short two-week intervention may account for the absence of compositional changes, highlighting the importance of longer supplementation periods such as those assessed in the present study. While the recent *in-vitro* TIM-2 experiment also demonstrated a modest butyrate increase (31), no statistically significant increase in fecal butyrate (despite a clear 30% rise) was observed in the pilot study by Pham et al. (32). In that exploratory study, the lack of significance could be due to the limited power from only 12 participants per arm. Furthermore, we hypothesize that elevated butyrate levels in the current trial were not maintained through week 12 may reflect increased microbial utilization, greater host absorption, shifts in community composition, or altered substrate availability, particularly in the context of a potential rise in total microbial biomass. In addition, fecal butyrate analysis is subject to high variability due to sample handling, transit time, absorption, and microbial cross-feeding (73).

As an exploratory analysis, we examined the distribution of riboflavin prototrophic and auxotrophic taxa using a metabolic modeling approach. Although the modeled species represented only about one-third of the total number of taxa (due to limitations in name-matching and the lack of metabolic models for all species), they jointly accounted for about 75% of total community abundance. Within this framework, we observed a notable increase in riboflavin-prototrophic species in the 10 mg group at week 12. Interestingly, this pattern is consistent with the significant enrichment of KEGG modules related to riboflavin biosynthesis at the same time and dose (Fig. 6 and 7). One explanation may be that increased extracellular riboflavin availability allows prototrophic bacteria to downregulate *de-novo* synthesis of riboflavin from GTP via feedback inhibition (left branch of pathway M00125 in Fig. 7C), thereby reducing metabolic burden and supporting growth. The resulting biomass increase of riboflavin-prototrophic bacteria, which continue converting extracellular riboflavin after uptake into FMN and FAD (right branch of pathway M00125 in Fig. 7C), could then account for the overall upregulation of the riboflavin synthesis pathway M00125. In fact, Thiele and colleagues systematically analyzed the genomes of 256 prevalent human gut bacteria and found riboflavin biosynthesis pathways in ∼50% of *Bacillota*, all *Bacteroidota* and *Fusobacteriota*, ∼92% of *Pseudomonadota*, and only two *Actinomycetota* genomes (20). In line with these observations, we detected characteristic increases in the relative abundance of *Bacillota* and *Bacteroidota*. Another explanation is that riboflavin prototrophs may benefit from cross-feeding with metabolically active riboflavin auxotrophs that indirectly support their expansion. Consistent with this, our metabolic modeling identified recurrent *Lachnospiraceae* taxa such as *Blautia_A_sp900066335* and the acetate- and butyrate-producing *Holdemanella biformis* (formerly *Eubacterium biforme*) as riboflavin auxotrophs. Additional auxotrophic taxa not captured by our modeling approach may further contribute.

We also found that the HACK Index, designed to reflect resilience and association with beneficial keystone taxa, significantly increased over 12 weeks in the 10 mg riboflavin group, suggesting enhanced microbial robustness upon colon-delivered riboflavin at this dosage. Moreover, the Dysbiosis Index showed a significant decrease with the 1.4 mg group, indicating a potential for microbial rebalancing even at low doses. Our network analyses revealed further dose-specific effects with 1.4 mg riboflavin inducing early changes in network modularity and significant increases in edge density and natural connectivity at week 4, indicative of a transient rise in microbial cohesion. When investigating dynamic changes over time, the 75 mg group demonstrated the greatest persistence and the strongest recruitment of new keystone taxa, potentially reflecting progressive network stabilization. In contrast, the placebo group showed limited overlap of keystone taxa across timepoints and a decline in unique keystone taxa, indicative of greater variability and reduced network stability. The 1.4 mg and 10 mg groups fell in between, with somewhat higher numbers of shared keystone taxa than placebo and evidence of taxa being gained or lost over time, suggesting dynamic restructuring and ecological adaptation toward a more stable network. Notably, the 10 mg dose yielded the highest number of unique keystone taxa by week 12, pointing to enhanced keystone recruitment and potentially reflecting optimal ecological remodeling. Together, these findings may highlight the complexity of riboflavin’s impact, with 1.4 mg possibly initiating early changes and microbiome rebalancing, 10 mg supporting optimal microbial innovation and resilience, and 75 mg favoring consolidation of an established network. However, these mechanisms remain difficult to disentangle, and future mechanistic studies will be required to clarify these subtle differences.

The functional KEGG module analysis provided further insight into microbial metabolic shifts. Although no between-group differences were significant, within-group comparisons revealed a dose-dependent increase in functional reprogramming, with 6, 51, and 96 significantly altered modules at 1.4, 10, and 75 mg of riboflavin respectively. The inclusion of the riboflavin biosynthesis module among the most significantly altered pathways emphasizes that riboflavin supplementation can engage and reshape microbial vitamin metabolism as discussed above. Importantly, the greatest functional response was observed at the 75 mg dose; however, this did not coincide with the most pronounced microbiome alterations in terms of taxonomic abundance, diversity, dysbiosis, or Hack index. This suggests that compositional changes in the microbiome do not directly translate into functional outcomes at this higher dose. Moreover, the early increase in butyrate could not be explained by the functional KEGG profile.

It should be considered that, beyond their roles in microbial cross-feeding, microbially produced or colon-delivered B-vitamins may also directly support host physiology, including MAIT cell activation and systemic vitamin supply (28,74). While dietary absorption occurs mainly in the small intestine, colonocytes express transporters for several B-vitamins, suggesting colonic uptake can contribute to systemic vitamin homeostasis, especially during insufficiency (20,75,76). Consistent with this, we observed a rise in plasma riboflavin from baseline to week 12 after 75 mg of colon-delivered riboflavin, confirming effective colonic absorption, in line with previously reported data (32).

In terms of clinical endpoints, we did not observe any significant effects on quality-of-life scores or gastrointestinal symptoms compared with placebo in this healthy study population. This suggests that, while certain microbiome and metabolic endpoints can shift, self-reported well-being and GI function remain largely unaffected in healthy adults, highlighting the need for studies in populations with greater potential for improvement.

The product was safe and well tolerated across all doses with no product-related SAEs or AEs and stable GSRS scores, supporting a favorable GI tolerability profile. Among systemic biomarkers, hs-CRP remained within the normal range across all groups while free thiols increased significantly within the 1.4 and 75 mg groups, indicating enhanced antioxidant capacity and reduced oxidative stress. Despite the lack of significance relative to placebo, the observation is consistent with previous reports in Crohn’s disease patients receiving 100 mg riboflavin daily for three weeks (27). Plasma sCD14 declined in all groups except for a modest rise at 75 mg riboflavin, which was not significant versus placebo, smaller than the placebo change, and remained below the physiological range (1500–2000 ng/mL), suggesting limited importance. Also, the within-group increases in fecal calprotectin at 10 and 75 mg riboflavin remained below the clinical threshold of 50 µg/g (77), with week 12 values comparable to, or lower than, placebo, indicating no clinical relevance. In addition, all reported AEs including calprotectin were judged unrelated to the study product and were evenly distributed across treatment and the placebo groups (Supplementary Figure S2).

A number of limitations warrant consideration. One is the use of microcrystalline cellulose as a placebo, which, although not a true prebiotic and used in small amounts, may still modestly modulate the microbiome and partly explain changes in the placebo group. Another limitation is that the cohort consisted of healthy individuals with clinical readouts and systemic/fecal biomarkers in normal ranges, leaving little room for improvement in clinical or symptom scores, a ceiling effect that may account for the absence of quality-of-life or gastrointestinal benefits despite detectable microbiome and metabolic shifts. Finally, future studies could employ absolute metagenomics, despite current challenges with variable fecal biomass and consistency, to determine whether taxa such as the dominant butyrate producers such as *Faecalibacterium* truly decline or remain stable relative to total microbial load, as recently suggested (78).

## Conclusion and outlook

Collectively, the results suggest that colon-delivered riboflavin influences the gut microbiota in a nuanced, dose-specific fashion, altering taxonomic and functional composition, improving network cohesion, influencing keystone taxa and enhancing metabolic potential including SCFA production. An optimal dose of about 10□mg emerged as the most effective evidenced by increases of alpha diversity, total species counts, KEGG module abundance including riboflavin synthesis, riboflavin prototrophic enrichment, and ecosystem stability. While 1.4□mg may trigger early but modest beneficial effects on modular network structure and performance, the 75□mg dose appears to drive broader functional but potentially weaker and less targeted taxonomical microbial responses. Although no effects were observed on clinical outcomes, the findings underscore the potential health-promoting potential of colon-targeted riboflavin, showing that precise delivery of riboflavin to the colon can enhance gut microbial function. This positions colon-targeted riboflavin as a functional microbiome modulator with potential in precision nutrition and microbiome-based therapies. To optimize colonic riboflavin supplementation, future trials could include microbiome pre-screening to quantify total riboflavin biosynthetic potential. Such targeted designs may help restore disrupted cross-feeding networks, thereby enhancing the colonization, activity, and metabolic output of key acetate and butyrate producing taxa central to maintaining intestinal health. Given that many acetate and butyrate-producing gut bacteria are in fact critically dependent on B vitamins for growth and metabolic activity (72,79), riboflavin may act as key metabolic ‘currency’ facilitating mutualistic exchanges between riboflavin-producing prototrophs and auxotrophs. Given that microbial B-vitamin synthesis is influenced by environment, lifestyle and diet (80,81), and colonic riboflavin deficiency has been linked to oxidative stress, insulin resistance, and intestinal inflammation (25,52,82,83), future research should target at-risk groups (IBS, IBD, or individuals with early metabolic or inflammatory markers), where detectable microbiome and clinical shifts are more likely.

## Data Availability

All data produced in the present study are available upon reasonable request to the authors

## Acknowledgements

We would like to thank Atlantia Clinical Trials (Cork, Ireland) for the excellent coordination and execution of this study, as well as for the fruitful discussions about the results obtained. We also acknowledge CMBio (Vedbæk, Denmark) for all metagenomic data processing, functional profiling and related statistical analysis. We thank Mick Watson and Alessio Milanese for data processing.

## Author’s contributions

RES, AR, WS, JWS conceptualized the study and interpreted the data. RES and WS drafted the manuscript. AM, CP, TA, PNM, EVLT, LW, FKO and JW performed the analysis and evaluation of microbiome-related data. All authors reviewed and approved the manuscript.

## Declarations

## Ethics approval and consent to participate

The study protocol, informed consent form, and all accompanying documents were reviewed and approved by the Clinical Research Ethics Committee of the Cork Teaching Hospitals (ECM 4(JJ) 22 February, 2022, Lancaster Hall, 6 Little Hanover Street, Cork, Ireland). The trial was conducted in full accordance with the ethical principles outlined in the Declaration of Helsinki (7th revision, October 2013), the International Council for Harmonisation (ICH) E6(R2) Good Clinical Practice (GCP) guidelines (November 2016), and all applicable local regulatory and legal requirements. Prior to initiation of any study-related procedures, written informed consent was obtained from all participants at the screening visit. The study was registered at clinicaltrials.gov (ID:NCT05803811, registration date 2023-02-16).

## Data availability

All data are available upon reasonable request from the corresponding author.

## Consent for publication

Not applicable.

