## Supplementary Information for "Colon-delivered vitamin B2 as a functional modulator of the human gut microbiome"

1. Health, Nutrition & Care (HNC), dsm-firmenich, Kaiseraugst, Switzerland.

2. Adelaide Medical School, Faculty of Health and Medical Sciences, CRE in Translating Nutritional Science to Good Health, The University of Adelaide. Australia

3. Microbiome Solutions, Niederhünigen, Switzerland.

4. Data Science & AI, Science and Research, dsm-firmenich, Delft, The Netherlands

5. Delft Bioinformatics Lab, Delft University of Technology, Delft, The Netherlands

6. Cmbio, Copenhagen, Denmark.

7. Data2time Sàrl, Geneva, Switzerland.

Keywords: Colonic Delivery, Riboflavin, Microbiome modulation, Butyrate, SCFA

*Corresponding author: Robert E. Steinert, Health, Nutrition & Care (HNC), dsm-firmenich, Wurmisweg 576, 4303 Kaiseraugst, Switzerland.

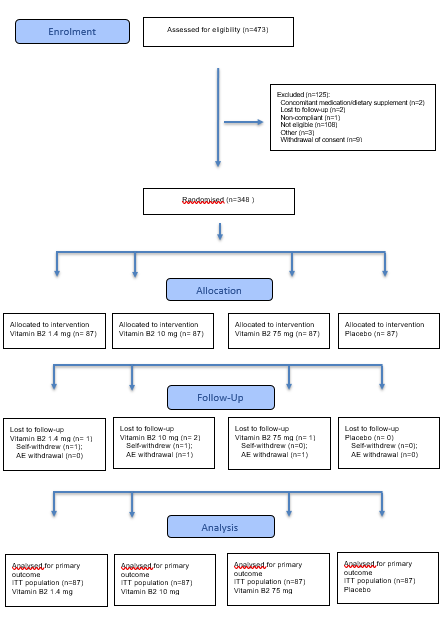

**Supplementary Figure S1 Effects of colon-delivered riboflavin on fecal microbial composition within groups.** CONSORT 2025 Flow Diagram depicting the progress through the phases of the randomized trial including enrolment, intervention allocation, follow-up, and data analysis

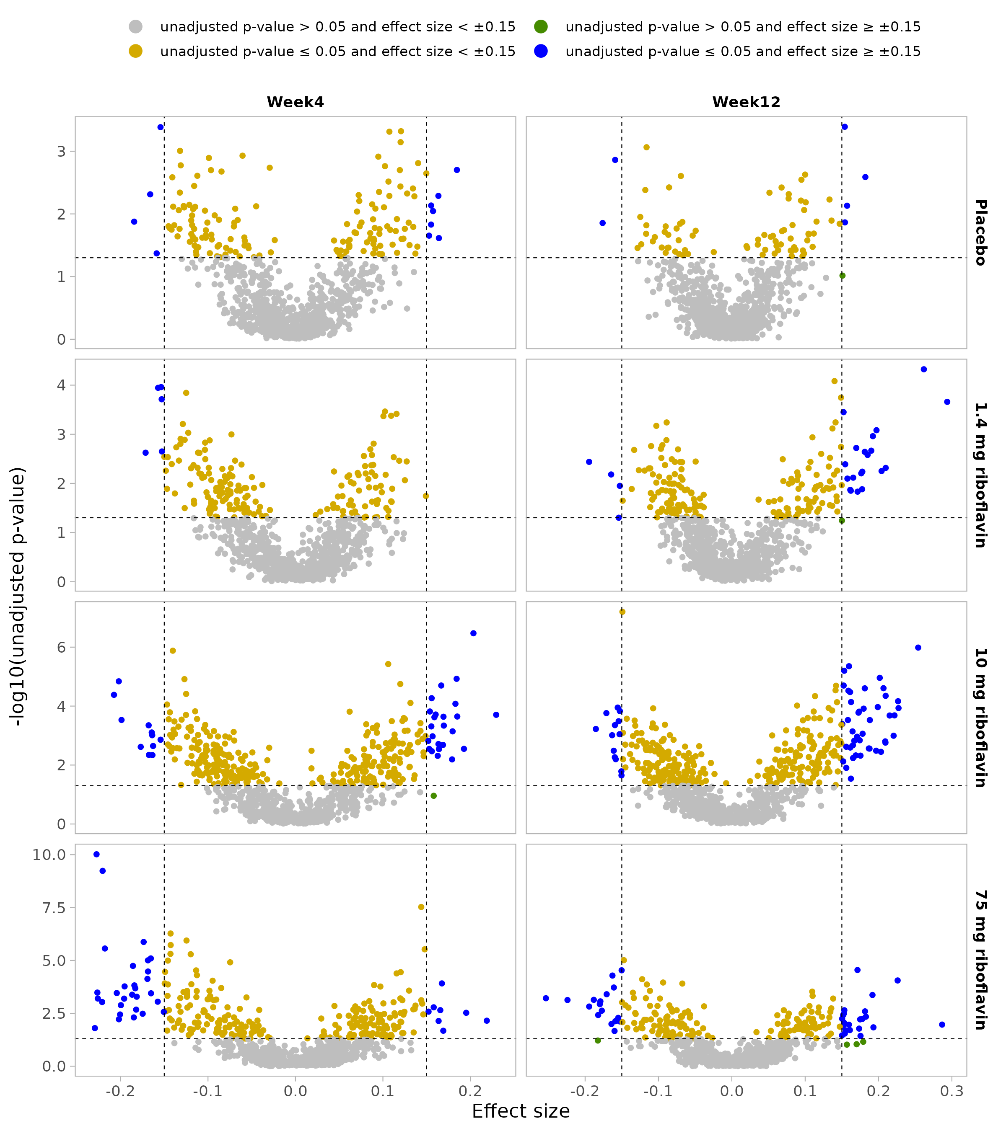

**Supplementary Figure S2 Effects of colon-delivered riboflavin on fecal microbial composition within groups**. Volcano plots illustrate the within-group differential abundant species at week 4 and week 12 when compared to baseline. Grey dots: unadjusted *p-*value > 0.05 and effect size < +/- 0.15; Orange dots: unadjusted *p-*value ≤ 0.05 and effect size < +/- 0.15; Green dots: unadjusted *p-*value > 0.05 and effect size ≥ +/- 0.15; Blue dots: unadjusted *p-*value ≤ 0.05 and effect size ≥ +/- 0.15.

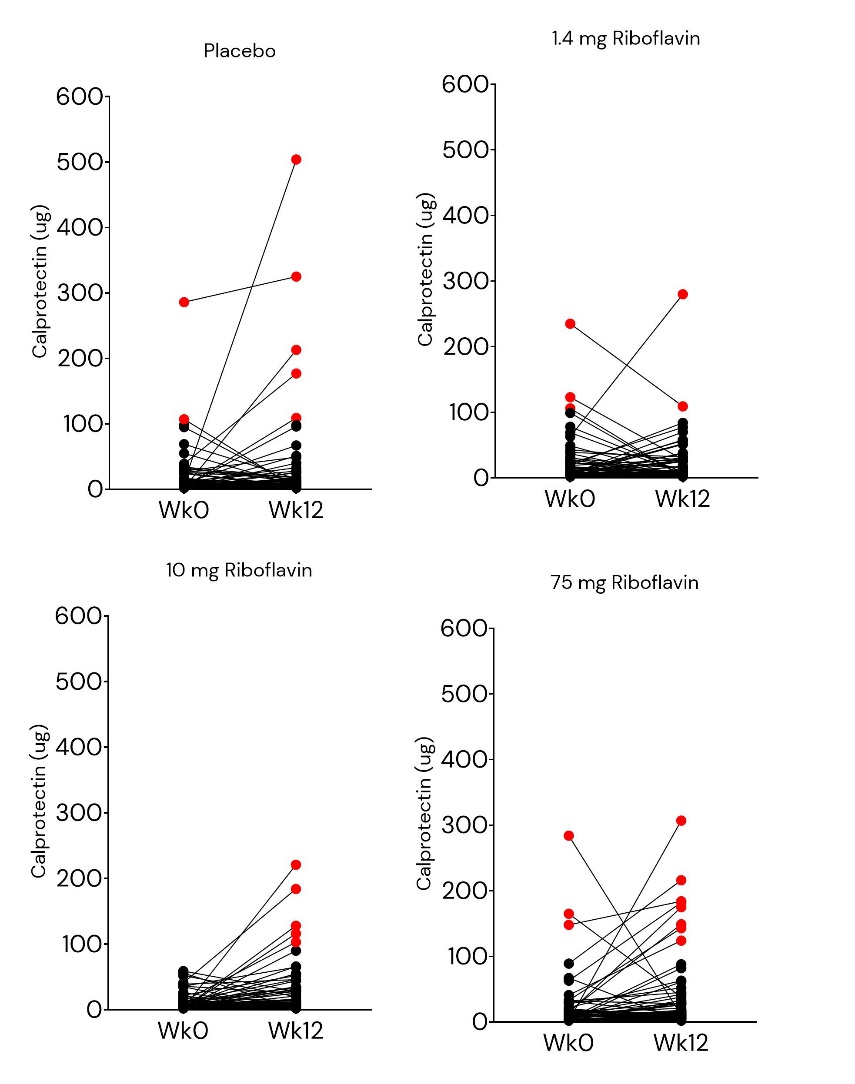

**Supplementary Figure S3** **Effects of colon-delivered riboflavin on fecal calprotectin.** Data represent individual fecal calprotectin concentrations (µg) at baseline and after 12 weeks in participants receiving placebo, 1.4 mg riboflavin, 10 mg riboflavin, or 75 mg riboflavin. Each line connects paired values from the same participant. Red dots indicate values above the clinical threshold of 100 µg.

**Supplemental Table S1 Baseline characteristics of the ITT populations**

|  | **Riboflavin  1.4 mg** | | **Riboflavin  10 mg** | | **Riboflavin  75 mg** | | **Placebo** | |
| --- | --- | --- | --- | --- | --- | --- | --- | --- |
|  | N | % | N | % | N | % | N | % |
| Sex – Female | 67 | 77.0 | 67 | 77.0 | 66 | 75.9 | 67 | 77.0 |
| Sex – Male | 20 | 23.0 | 20 | 23.0 | 21 | 24.1 | 20 | 23.0 |
| Age | 57.66 (5.65) | | 57.68 (5.65) | | 58.18 (6.24) | | 57.68 (5.90) | |
| Weight | 70.63 (11.19) | | 70.61 (11.66) | | 69.89 (10.79) | | 70.39 (10.74) | |
| BMI | 25.19 (2.76) | | 25.30 (2.63) | | 25.18 (2.67) | | 25.05 (2.64) | |

Age, weight and BMI, values are shown as mean (SD)

**Supplemental Table S2** **Effects of colon-delivered riboflavin on fecal microbial composition between groups**

Data represent between-group differential abundant OTUs at species level at week 4 and week 12 when compared to placebo. with a cut-off in effect of ± 0.15 and a *p-*value <0.05. Tables are sorted by effect size. Table complements Fig. 1. shown in main document. (Note: In this Table the old phylum names are still in use: *Proteobacteria* (new *Pseudomonadota*), *Firmicutes* (new *Bacillota*), *Bacteroidetes* (new *Bacteroidota*). See xls file *Supp Table S2*

**Supplemental Table S3** **Effects of colon-delivered riboflavin on fecal microbial composition within groups**

Data represent within group-group differential abundant OTUs at species level at week 4 and week 12 when compared to baseline placebo. with a cut-off in effect of ± 0.15 and a p-value <0.05. Tables are sorted by effect size. Table complements Supplementary Figure S1. (Note: In this Table the old phylum names are still in use: *Proteobacteria* (new *Pseudomonadota*), *Firmicutes* (new *Bacillota*), *Bacteroidetes* (new *Bacteroidota*). See xls file *Supp Table S3*

**Table S4** **Effects of colon-delivered riboflavin on modular network structure** **and performance**

| **Network** | **Node** | **Edge density** | **Clustering coefficient** | **Modularity** | **Positive edge percentage** | **Natural connectivity** |
| --- | --- | --- | --- | --- | --- | --- |
| Week 0 |  |  |  |  |  |  |
| Placebo | 761 | 0.01644 | 0.08815 | 0.38126 | 71.37148 | 0.00220 |
| Vit B2 1.4 mg | 761 | 0.01690 | 0.08909 | 0.38153 | 70.32337 | 0.00222 |
| Vit B2 10 mg | 761 | 0.01610 | 0.08865 | 0.38481 | 69.59416 | 0.00218 |
| Vit B2 75 mg | 761 | 0.01593 | 0.08777 | 0.38968 | 72.16674 | 0.00217 |
| Week 4 |  |  |  |  |  |  |
| Placebo | 761 | 0.01597 | 0.09400 | 0.38890 | 73.04028 | 0.00217 |
| Vit B2 1.4 mg | 761 | **0.01673*** | 0.08764 | 0.39975 | 71.82138 | **0.00222**** |
| Vit B2 10 mg | 761 | 0.01624 | 0.09400 | 0.38767 | 73.04028 | 0.00220 |
| Vit B2 75 mg | 761 | 0.01637 | 0.08973 | 0.40628 | 71.82087 | 0.00220 |
| Week 12 |  |  |  |  |  |  |
| Placebo | 761 | 0.01628 | 0.08865 | 0.37811 | 71.52262 | 0.00219 |
| Vit B2 1.4 mg | 761 | 0.01690 | 0.08981 | 0.37171 | 69.89357 | 0.00223 |
| Vit B2 10 mg | 761 | 0.01650 | 0.08952 | 0.39179 | 69.94971 | 0.00221 |
| Vit B2 75 mg | 761 | 0.01637 | 0.08999 | 0.38077 | 70.54722 | 0.00220 |

Data represent network properties across the 12 constructed networks, grouped by time point and treatment group. At each time point, the networks constructed for each riboflavin dose were compared with the network constructed for the placebo group. ** = *p < 0.01*, * = *p < 0.05*.

**Supplemental Table S5** **Individual species ability to grow with and without riboflavin based on available metabolic models**

Data show 245 species and their trophy status with 86 species being considered riboflavin prototroph and 159 species riboflavin auxotroph. Models of these species were identified by name matching to AGORA2 and genome-based matching to APOLLO. See xls file *Supp Table S5*
